# Interpretable Transformer-Based Phase Recognition for Transabdominal Preperitoneal Laparoscopic Inguinal Hernia Repair

**DOI:** 10.64898/2026.04.26.26351777

**Authors:** Marzie Lafouti, Liane S. Feldman, Amir Hooshiar

**Affiliations:** Surgical Performance Enhancement and Robotics Centre (SuPER), Department of Surgery, McGill University, Montréal, QC, Canada

**Keywords:** Surgical phase recognition, TAPP laparoscopic inguinal hernia repair, Vision transformers, SurgFormer, Context-aware operating room

## Abstract

**Background:** Surgical phase recognition is a critical prerequisite for context-aware operating rooms and automated skill assessment. While artificial intelligence (AI) benchmarking has progressed for simpler procedures, applying surgical phase recognition to complex, anatomically demanding operations like transabdominal preperitoneal (TAPP) laparoscopic inguinal hernia repair (LIHR) remains uncharted, limiting the scalability of AI-driven analysis in this globally frequent surgery.

**Methods:** We introduced a workflow analysis framework for TAPP utilizing SurgFormer, a vision transformer architecture. The model was evaluated on an institutional dataset annotated via the Theator platform, with ethical approval from the Research Ethics Board (REB) of the McGill University Health Centre (MUHC). To mitigate data scarcity, we employed a three-stage sequential transfer learning strategy, leveraging weights from Kinetics-400 and Cholec80 before domain adaptation to visual complexities of LIHR.

**Results:** The framework achieved a peak Top-1 accuracy of 90.64% through a cumulative training approach with 22 videos, outperforming standard full-set fine-tuning. Beyond predictive metrics, dimensionality reduction and embedding analysis (PCA, t-SNE, and UMAP) across the model’s attention blocks revealed a maturation of internal representations, evolving from local textures to distinct, high-level semantic surgical phases.

**Conclusion:** This study presents a novel, highly accurate application of transformer-based surgical phase recognition to TAPP. By mapping the intricate LIHR workflow and providing deep interpretability, this work establishes a foundation for real-time intraoperative guidance and objective performance profiling in hernia surgery.

## 1. Introduction

### 1.1. Background and clinical need

The modern operating room is undergoing a profound paradigm shift towards the context-aware operating room [15]. At the heart of this transformation is surgical phase recognition, which is the ability of an AI system to automatically identify, track, and contextualize the discrete steps of a surgical procedure in real-time. Surgical phase recognition is not merely an observational or administrative tool; it is the foundational building block for a host of advanced, next-generation clinical applications [15, 9]. Accurate, real-time workflow mapping enables objective and automated surgical skill assessment, dynamic estimation of remaining operative time for optimized hospital resource management, and the automated indexing of massive surgical video archives for targeted resident training [5]. Ultimately, robust surgical phase recognition is a necessary precursor to intraoperative cognitive support systems, which aim to warn surgeons of impending anatomical hazards or deviations from the standard of care before adverse events occur.

Despite its vast potential and the rapid acceleration of AI in healthcare, the development of clinical-grade surgical phase recognition systems has been heavily skewed towards relatively linear and standardized procedures, most notably laparoscopic cholecystectomy. Conversely, laparoscopic inguinal hernia Repair (LIHR), particularly the transabdominal preperitoneal (TAPP) approach, remains critically under-explored in the domain of surgical data science [6, 3]. TAPP is one of the most frequently performed minimally invasive surgeries worldwide, with over 20 million LIHRs performed globally each year [19]. Yet, it presents a uniquely challenging workflow, characterized by complex preperitoneal anatomy, subtle visual transitions, and significant tool-tissue occlusion that pushes the limits of current computer vision models [20, 23].

The procedure requires meticulous navigation through delicate anatomical planes, specifically the dissection of the preperitoneal space (including the Bogros and Retzius spaces), the careful reduction of the hernia sac, the precise manipulation and fixation of a prosthetic mesh, and complex suturing for peritoneal closure [25, 20]. The high inter-surgeon variability in handling these steps, combined with frequent instrument-tissue occlusions and subtle visual transitions between phases, creates a highly complex visual landscape [20]. Furthermore, while extensive, heavily annotated public datasets exist for cholecystectomy (such as Cholec80 [22]), robustly annotated multi-phase data for TAPP is remarkably scarce [6]. This data bottleneck has severely limited the development, training, and validation of tailored AI models capable of decoding the complex spatial-temporal narrative of hernia repair.

### 1.2. State-of-the-art (SOTA)

The automated analysis of surgical workflows has evolved significantly over the past decade, mirroring the broader advancements in computer vision and deep learning. Early approaches to Surgical Phase Recognition relied heavily on combining Convolutional Neural Networks (CNNs) for spatial feature extraction with traditional machine learning classifiers or Hidden Markov Models (HMMs). A landmark advancement in this era was the introduction of EndoNet [22], which successfully utilized a standard CNN architecture to extract visual features from laparoscopic videos for phase and tool recognition. While effective for localized, frame-by-frame analysis, these early spatial architectures suffered from significant temporal instability, often resulting in erratic phase predictions and “flickering” due to their inability to contextualize the current frame within the broader surgical timeline.

To address these limitations, the field transitioned toward temporal modeling techniques. Researchers began pairing CNN-extracted spatial features with Recurrent Neural Networks (RNNs), specifically Long Short-Term Memory (LSTM) networks, such as in the SV-RCNet architecture [8]. Subsequently, Temporal Convolutional Networks (TCNs), like the TeCNO model [4], gained prominence for their ability to model longer temporal windows using dilated convolutions. While these temporal smoothing techniques successfully reduced prediction flickering and improved overall accuracy, they inherently struggle to capture long-range temporal dependencies. In complex procedures like TAPP, an event occurring at minute 10 (e.g., the extent of peritoneal dissection) directly influences the visual context at minute 40 (e.g., the specific technique required for mesh placement). RNNs and TCNs often fail to retain this distant global context [4, 14].

Recently, vision transformers (ViTs) have emerged as the state-of-the-art in video understanding [1, 18]. By abandoning recurrence and convolution in favor of self-attention mechanisms, transformers can model complex spatial-temporal redundancies by processing sparse frames and establishing true global context. Foundational models like TimeSformer [1] proved the efficacy of divided space-time attention in generic video datasets like Kinetics-400. In the surgical domain, architectures such as SurgFormer [24] have successfully adapted these concepts, demonstrating superior capabilities in modeling complex surgical workflows by leveraging hierarchical temporal attention to track long-range procedural steps [24, 17].

Parallel to these evolutions in workflow-level analysis, significant progress has been made in extracting granular, highly accurate data from the surgical scene. Early efforts in visual object tracking focused on instrument localization in specialized procedures, such as capsulorhexis eye surgery, utilizing Siamese networks [10]. More recently, these tracking capabilities have been adapted to the complex laparoscopic environment, demonstrating the feasibility of robust, real-time instrument tracking and segmentation through deep fusion methodologies [11, 16] and natural language-guided scene inference [7]. Building upon this foundational spatial understanding, recent studies have also explored the integration of interpretable spatiotemporal AI pipelines to accelerate objective surgical skill assessment via saliency-guided highlight generation [12, 13].

However, despite these strides in object-level tracking and highlight generation, a critical gap remains: adapting state-of-the-art sequential transformer architectures to accurately map the entire procedural continuum of a highly complex, multi-step surgery like TAPP. Furthermore, as deep learning models increasingly permeate the surgical domain, the “black box” nature of these algorithms poses a significant barrier to clinical trust and adoption. There is a pressing need not only to achieve high predictive accuracy on scarce surgical datasets but also to transparently visualize exactly how these complex transformer models learn, represent, and differentiate complex surgical concepts.

### 1.3. Objectives and contributions

To bridge the gap between advanced deep learning capabilities and the clinical complexities of hernia repair, this study presents a novel, highly interpretable application of the SurgFormer architecture for robust phase recognition in TAPP. Utilizing a proprietary, REB-approved dataset of surgical videos obtained from the McGill University Health Centre (MUHC) and expertly annotated via the cloud-based Theator platform, we aim to map the intricate seven-phase workflow of LIHR. To overcome the inherent scarcity of annotated TAPP data, we employ a sequential domain-adaptation strategy, leveraging knowledge transferred from generic video (Kinetics-400) and benchmark surgical procedures (Cholec80) [2].

The primary contributions of this work are fourfold:

1. We introduce an accurate transformer-based surgical phase recognition frameworks tailored specifically for TAPP hernia repair. By fine-tuning a Kinetics-pretrained model first on Cholec80 and subsequently on our institutional TAPP dataset, we demonstrate that sequential knowledge transfer effectively mitigates data scarcity, achieving an 90.64% accuracy on unseen clinical data.
2. We benchmark the intermediate architecture on the Cholec80 dataset, achieving an 89.37% accuracy, thereby verifying the model’s baseline spatial-temporal modeling capabilities prior to transitioning to the complex hernia domain.
3. We develop continuous, progressive phase map visualizations integrated synchronously with the native 25 frames per second (fps) surgical video. This approach juxtaposes ground truth annotations with real-time predictions, clearly highlighting transient errors and the temporal evolution of surgical phases for intuitive clinical review.
4. To demystify the transformer’s decision-making process, we treat the fully trained SurgFormer as a feature extractor and conduct a granular dimensional reduction and embedding analysis (utilizing PCA, t-SNE, and UMAP) across all 12 of its attention blocks. This explicitly illustrates how the model’s internal representations mature progressively from parsing low-level visual textures in early layers to distinctly clustering complex, high-level semantic surgical phases in the final layers.

## 2. Methodology

### 2.1. Dataset architecture and privacy

To develop and validate the proposed phase recognition framework, this study utilized two distinct surgical video datasets: a public benchmark dataset for initial domain adaptation and a proprietary institutional dataset for target task fine-tuning. A comparison of the procedural phases is summarized in Table 1.

**Table 1:**
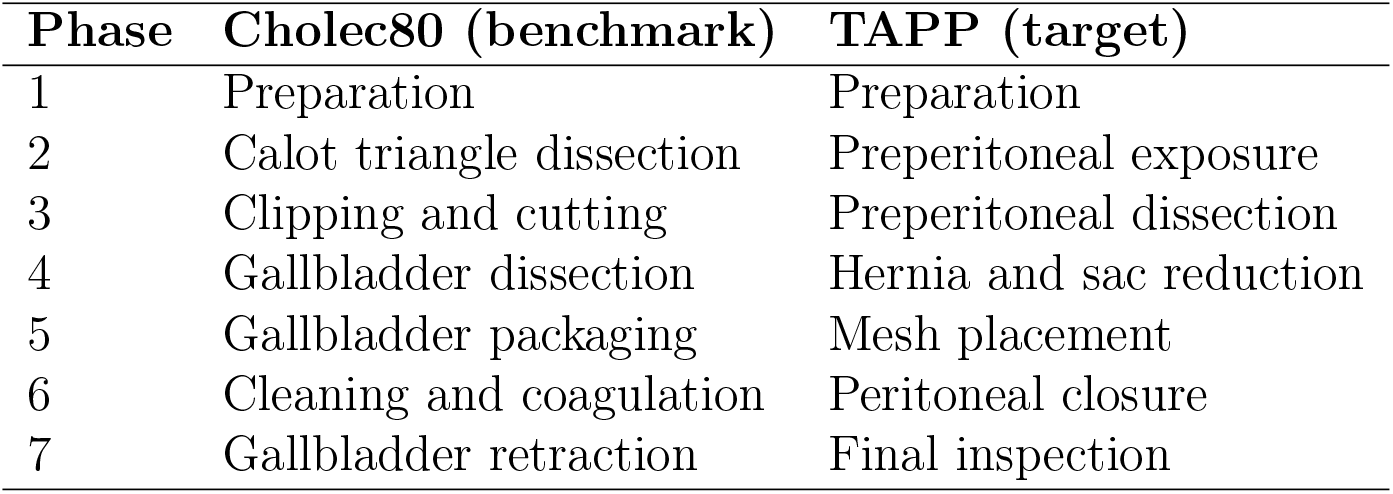
Comparison of cholecystectomy (Cholec80) and TAPP LIHR phases.

| Phase | Cholec80 (benchmark) | TAPP (target) |
| --- | --- | --- |
| 1 | Preparation | Preparation |
| 2 | Calot triangle dissection | Preperitoneal exposure |
| 3 | Clipping and cutting | Preperitoneal dissection |
| 4 | Gallbladder dissection | Hernia and sac reduction |
| 5 | Gallbladder packaging | Mesh placement |
| 6 | Cleaning and coagulation | Peritoneal closure |
| 7 | Gallbladder retraction | Final inspection |

#### 2.1.1. Institutional TAPP dataset

The primary dataset consists of 32 full-length videos of TAPP procedures performed at the McGill University Health Centre (MUHC). The videos were securely handled and annotated using the Theator surgical intelligence platform [21] in compliance with patient privacy protocols. The dataset was split into a training set of 25 videos (140,810 frames) and an unseen test set of 7 videos (24,628 frames) for final evaluation.

To standardize the surgical narrative and ensure clinical relevance, the procedure was segmented into seven distinct phases, adopted from the established definitions by Takeuchi et al. [20]: Preparation, preperitoneal exposure, preperitoneal dissection, hernia and sac reduction, mesh placement, peritoneal closure, and final inspection (Table 1).

#### 2.1.2. Cholec80 benchmark dataset

To bridge the domain gap between generic natural videos and the highly specialized hernia repair domain, we utilized the publicly available Cholec80 dataset [22] as an intermediate training step. Cholec80 consists of 80 laparoscopic cholecystectomy videos annotated with seven distinct phases. The dataset was divided into standard benchmark splits of 40 videos for training (1,527,708 frames) and 40 for testing (2,454,890 frames), with frames extracted at 1 fps.

### 2.2. Model architecture: SurgFormer

For spatial-temporal sequence modeling, we utilized the SurgFormer architecture [24]. Unlike traditional CNN-RNN pipelines, SurgFormer is a pure Vision Transformer (ViT) designed to capture long-range dependencies within surgical videos. The core of the architecture relies on a divided space-time attention mechanism, allowing the model to independently calculate spatial self-attention within individual frames and temporal self-attention across sequences of frames. This hierarchical approach effectively handles the visual redundancies and temporal continuity inherent in surgical workflows. The network consists of 12 sequential transformer blocks, which progressively process local visual features into high-level, global semantic representations of the surgical action.

### 2.3. Training strategy and experimental variants

Training deep transformer architectures from scratch on small medical datasets frequently leads to severe overfitting. To mitigate the scarcity of TAPP annotations, we implemented a sequential, three-stage transfer learning methodology:

1. **Base initialization:** The model’s initial weights were transferred from a TimeSformer [1] pre-trained on the Kinetics-400 dataset, a large-scale human action recognition database. This provided the network with foundational spatial-temporal feature extraction capabilities.
2. **Surgical domain adaptation:** The model was then fine-tuned on the Cholec80 training set to adapt its representations from generic human actions to the specific visual domain of laparoscopic surgery. This training was conducted over 50 epochs.
3. **Target task fine-tuning:** Finally, the best-performing Cholec80 checkpoint was utilized to initialize the model for the TAPP LIHR dataset. The network was fine-tuned on the TAPP training videos for 50 epochs. The best model weights were saved each epoch, where the minimum validation loss and optimal validation accuracy were observed, effectively preventing overfitting on the smaller dataset.

To further investigate the data efficiency and learning stability of the SurgFormer architecture, three training protocols were implemented, which are direct training, casecade training and cumulative training.

#### Direct training

The model was trained directly on the 25 TAPP videos for 50 epochs, initialized with TimeSformer weights (Kinetics-400), bypassing the Cholec80 domain adaptation step.

#### Cascade training

To assess incremental learning capabilities, the 25 training videos were divided into 13 sequential chunks (2 videos per chunk). The model was initialized with Cholec80 weights and fine-tuned on the first chunk.

Each subsequent training iteration utilized the weights from the previous iteration to train on the next 2-video chunk.

#### Cumulative training

The model (initialized with Cholec80 weights) was trained on increasingly larger subsets of the TAPP dataset to determine the minimum data threshold for high-accuracy recognition, starting with 2 videos and adding chunks of 2 until reaching the full 25-video set.

### 2.4. Evaluation and embedding analysis

Model performance was primarily evaluated using overall frame-wise accuracy and continuous phase map visualizations on the unseen test sets. To provide a qualitative assessment of temporal stability, we reconstructed the test predictions into full-resolution, 25 fps surgical videos. These visualizations juxtapose the ground truth phase, the predicted phase, and a progressive color-coded timeline highlighting transient classification errors in real-time.

Furthermore, to interpret the “black box” learning dynamics of the transformer, we conducted a progressive embedding analysis. The fully trained, TAPP fine-tuned SurgFormer was deployed as a feature extractor. Video clips were fed into the model, and the high-dimensional feature representations were extracted from the outputs of multiple stages across the 12 transformer blocks. To visualize these representations in 2D and 3D space, we applied three distinct dimensionality reduction techniques: Principal component analysis (PCA), t-distributed stochastic neighbor embedding (t-SNE), and uniform manifold approximation and projection (UMAP). This analysis aimed to explicitly trace how the model’s internal embeddings evolve from entangled, low-level local visual patterns in the early layers to distinctly clustered, high-level semantic surgical phases in the terminal layers.

## 3. Results

### Domain adaptation with SOTA

The initial phase of our sequential transfer learning strategy involved fine-tuning the Kinetics-pretrained model on the Cholec80 benchmark dataset. The SurgFormer architecture successfully adapted to the general laparoscopic visual domain, achieving an overall frame-wise accuracy of 89.37% on the Cholec80 test set (Table 2). The model demonstrated steady convergence during this intermediate training phase, with the optimal validation accuracy observed at epoch 20. This robust baseline confirmed the architecture’s capacity for complex spatial-temporal modeling prior to targeted hernia adaptation.

**Table 2:** Comparative performance of the training protocols: Zero-shot inference, direct, cascade and cumulative training.

| Strategy | Init. weights | Train set | Test set | Max acc. |
| --- | --- | --- | --- | --- |
| Direct | K-400 | 40 (Cholec80) | 40 (Cholec80) | 89.37% (Ep. 20) |
| Zero-shot | K-400 | 40 (Cholec80) | 7 (TAPP) | 15.77% |
| Direct | K-400 | 25 (TAPP) | 7 (TAPP) | 89.89% (Ep. 7) |
| Cascade | Cholec80 | Seq. chunks | 7 (TAPP) | 87.99% (It. 8) |
| Cumulative | Cholec80 | 25 (TAPP) | 7 (TAPP) | 89.99% (Ep. 18) |
| **Cumulative** | **Cholec80** | **22 (TAPP)** | **7 (TAPP)** | **90.64% (Ep. 10)** |

We evaluated the Cholec80-pretrained model directly on the unseen TAPP test set without any target domain adaptation (zero-shot transfer) to quantify the necessity of targeted fine-tuning. As detailed in Table 2, the zero-shot model achieved a Top-1 accuracy of only 15.77%. However, following fine-tuning on the TAPP dataset, the model’s performance improved dramatically, achieving a 90.64% Top-1 accuracy and an 86.44% Mean F1-Score. The detailed class-wise performance of the fine-tuned model using the three training strategies is visualized in the confusion matrices in Figure 1.

**Figure 1:**
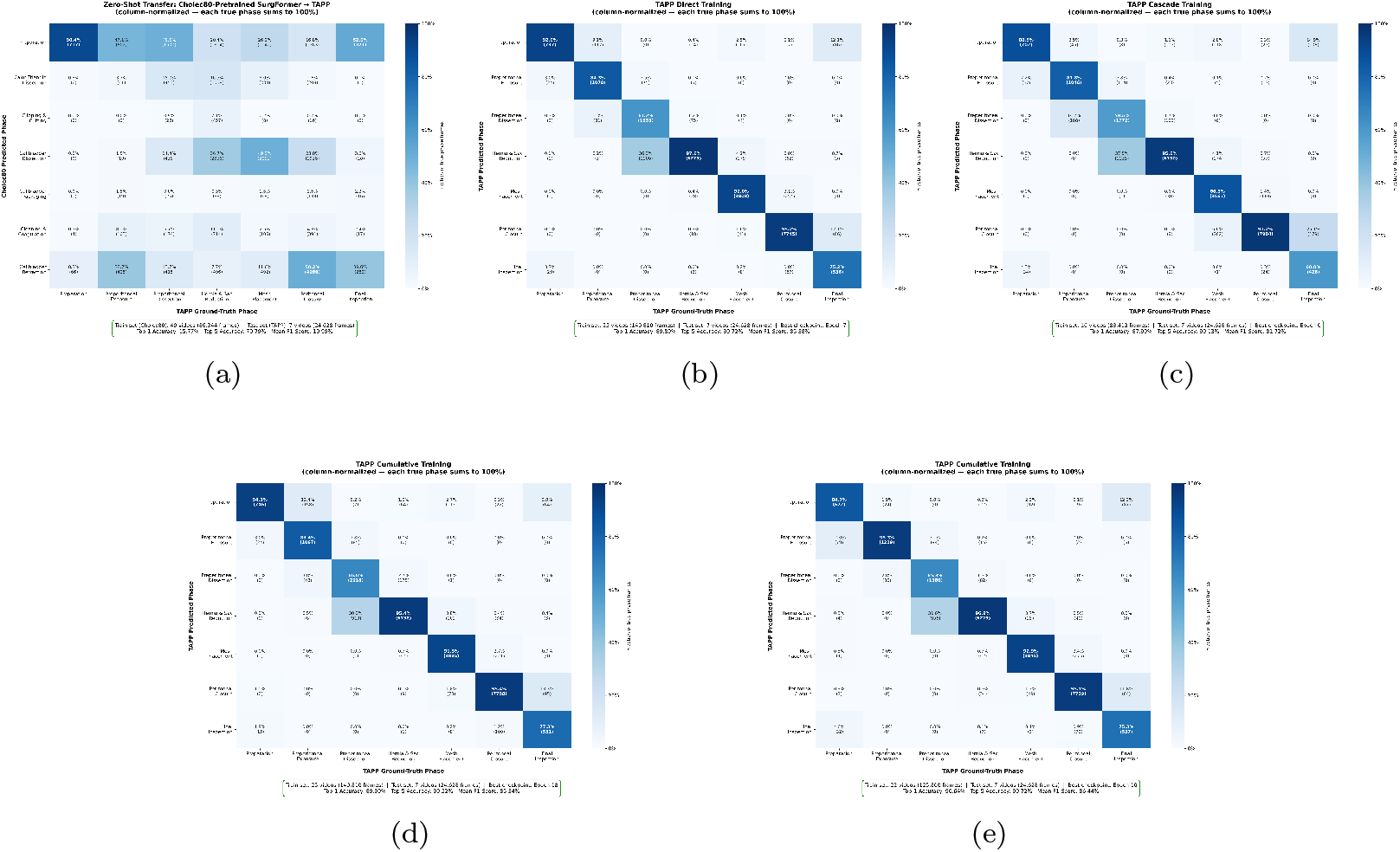
Comparison of normalized confusion matrices for TAPP phase recognition across training protocols: (a) Zero-shot transfer, (b) Direct training, (c) Cascade training (Iter. 8), (d) Cumulative training (25 videos), and (e) Optimal cumulative training (22 videos).

### Comparative analysis of training protocols

To identify the optimal fine-tuning strategy, we compared three training protocols: Direct, cascade, and cumulative training. The quantitative results are detailed in Table 2. The standard fine-tuning protocol on the full 25-video TAPP set yielded a robust accuracy of 89.99%. However, the cumulative training strategy demonstrated that peak performance could be achieved with a smaller data subset; specifically, with 22 training videos, the model reached its highest recorded Top-1 accuracy of 90.64% at epoch 10. The cascade training approach, which fine-tuned the model on sequential 2-video chunks, achieved its peak accuracy of 87.99% at the 8th iteration.

Furthermore, we analyzed performance as training volume increased from 2 to 25 videos to identify the optimal data threshold for TAPP phase recognition (Table 3). The results indicate a steady upward trend in accuracy as more procedural diversity is introduced, with performance peaking at 22 videos. Interestingly, the inclusion of the final three videos resulted in a marginal performance dip to 89.99%, suggesting that maximum generalizability is reached prior to utilizing the full dataset.

**Table 3:** Accuracy range across cumulative training iterations on TAPP.

| No. videos | Top-1 acc. | No. videos | Top-1 acc. |
| --- | --- | --- | --- |
| 2 | 81.81% | 14 | 88.85% |
| 4 | 87.66% | 16 | 88.59% |
| 6 | 87.38% | 18 | 88.93% |
| 8 | 87.80% | 20 | 89.55% |
| 10 | 89.10% | **22** | **90.64%** |
| 12 | 87.90% | 25 | 89.99% |

### Qualitative workflow visualization

To translate these quantitative metrics into clinically actionable insights, we generated continuous temporal visualizations of the model’s predictions. Figure 2 illustrates the color-coded phase progression for the TAPP test set, directly juxtaposing the ground truth annotations against the model’s predictions. Misclassified frames are explicitly highlighted in red, revealing that the majority of predictive errors are transient and highly localized to the subtle, transitional boundaries between consecutive surgical phases.

**Figure 2:**
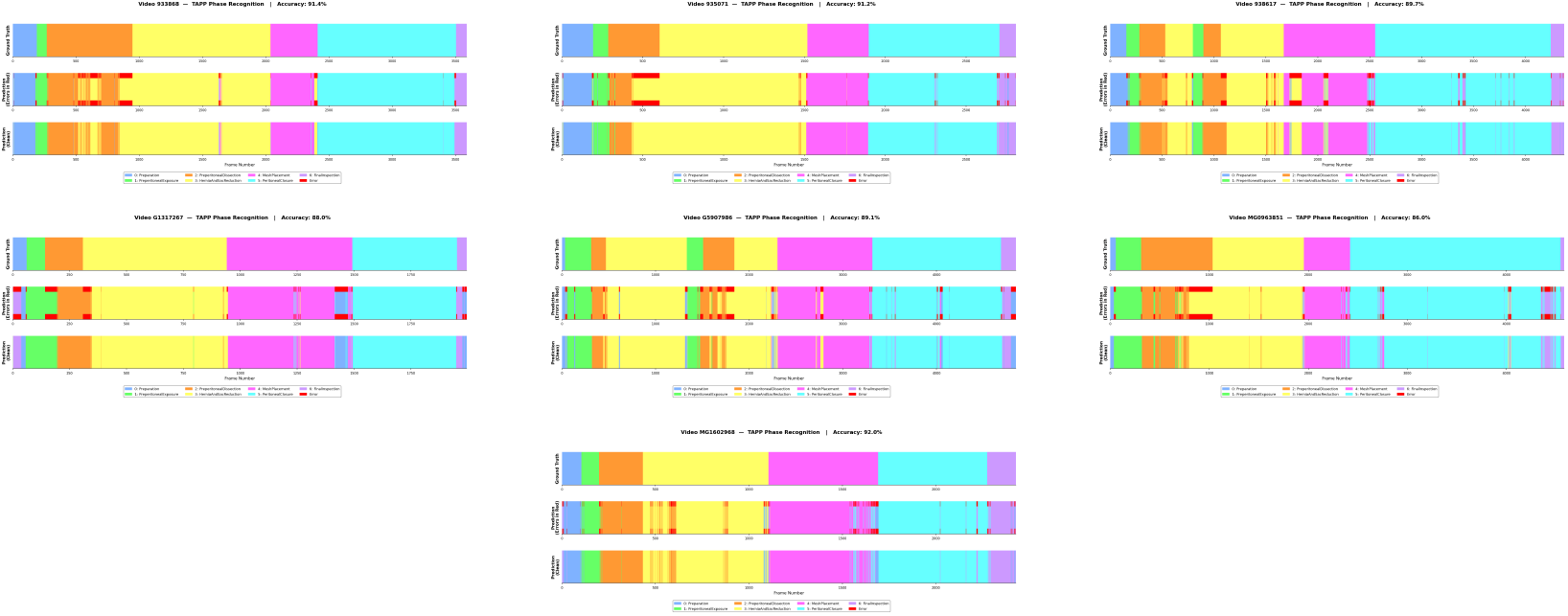
Comprehensive qualitative evaluation of automated TAPP phase recognition. This grid presents the progressive phase bars for the seven full-length, unseen test videos. Each sub-panel compares the clinician-annotated ground truth segmentation against the model’s generated predictions, with transient classification errors highlighted.

Furthermore, we developed a synchronous, dual-panel video playback system operating at the native 25 fps. As depicted in Figure 3, the system projects the real-time predicted phase and ground truth label directly onto the surgical feed (top-left), while simultaneously generating a progressive phase map (right panel). This dynamic interface allows clinicians to visually track the evolution of the surgery alongside the model’s real-time inference and transient error states.

**Figure 3:**
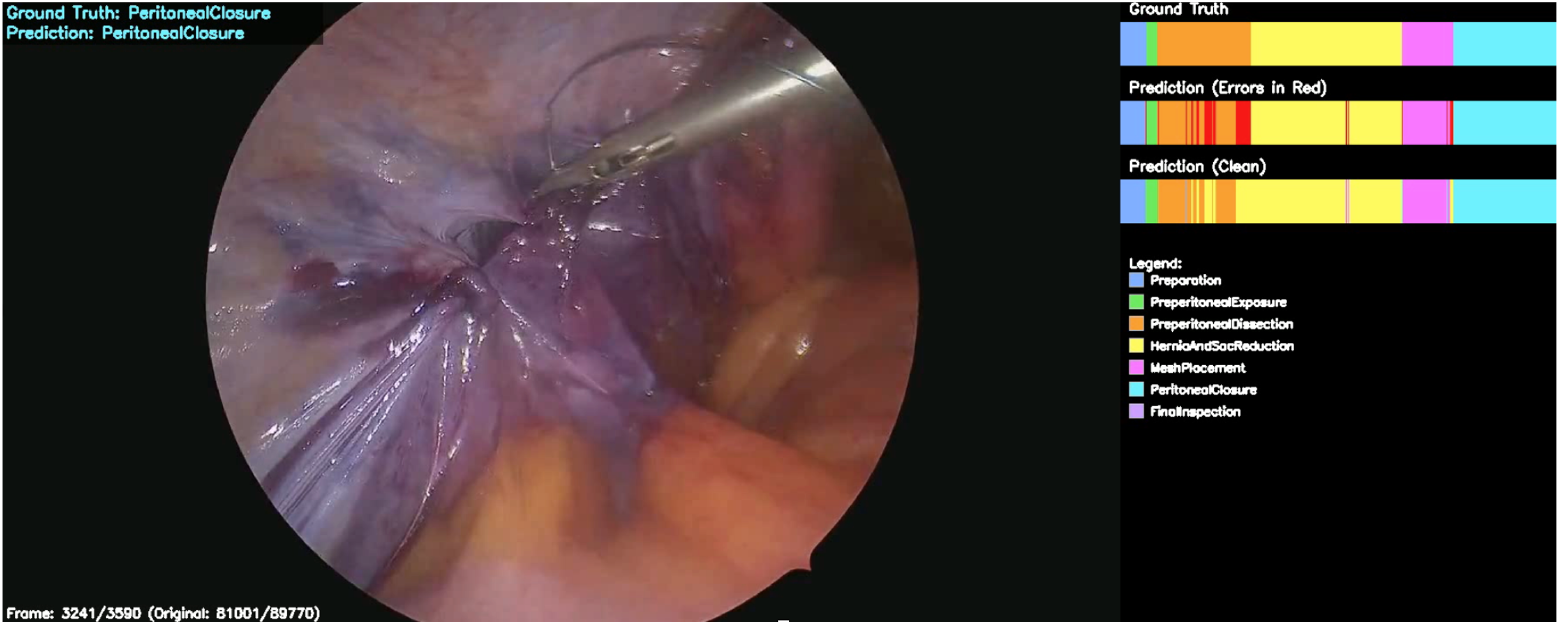
Final intraoperative phase recognition interface for LIHR. This integrated visualization tool combines the synchronous 25 fps surgical video stream (left) with key real-time overlays: (top-left) instantaneous predicted phase and ground truth label, and (right) a progressive, color-coded phase map that dynamically updates with predicted and ground truth timelines, explicitly marking misclassification events.

### Progressive embedding analysis

To interpret the internal learning dynamics of the transformer architecture, a comprehensive dimensionality reduction analysis was performed using PCA, t-SNE, and UMAP across the 12 attention blocks (Figure 4). The analysis reveals a distinct, progressive maturation of feature representations as data propagates through the network depth. In the initial blocks, the high-dimensional embeddings exhibit significant entanglement. However, in the terminal blocks, the representations resolve into highly distinct, separable clusters corresponding precisely to the 7 semantic surgical phases. Figures 5 and 6 illustrate the propagation of data through the depth of the network in two-dimensional and three-dimensional, respectively.

**Figure 4:**
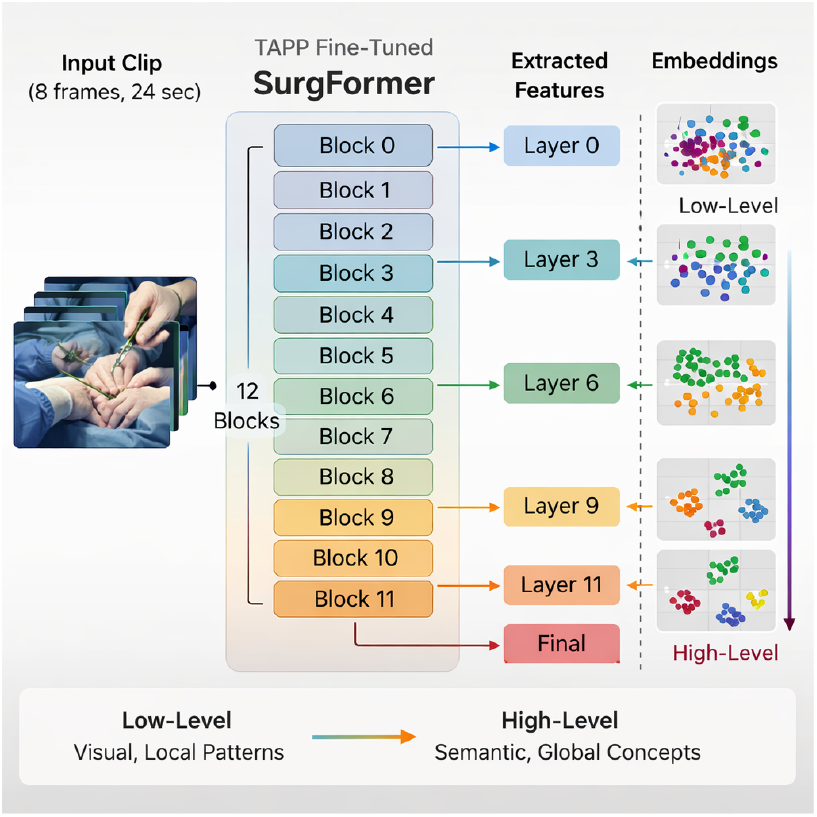
Overview of the progressive embedding extraction methodology. The fine-tuned SurgFormer architecture processes input video clips through 12 sequential transformer blocks. High-dimensional feature representations are systematically extracted from the outputs of intermediate layers (blocks 0, 3, 6, 9, 11, and the final layer) to trace the evolution from low-level visual patterns to high-level semantic phase clusters.

**Figure 5:**
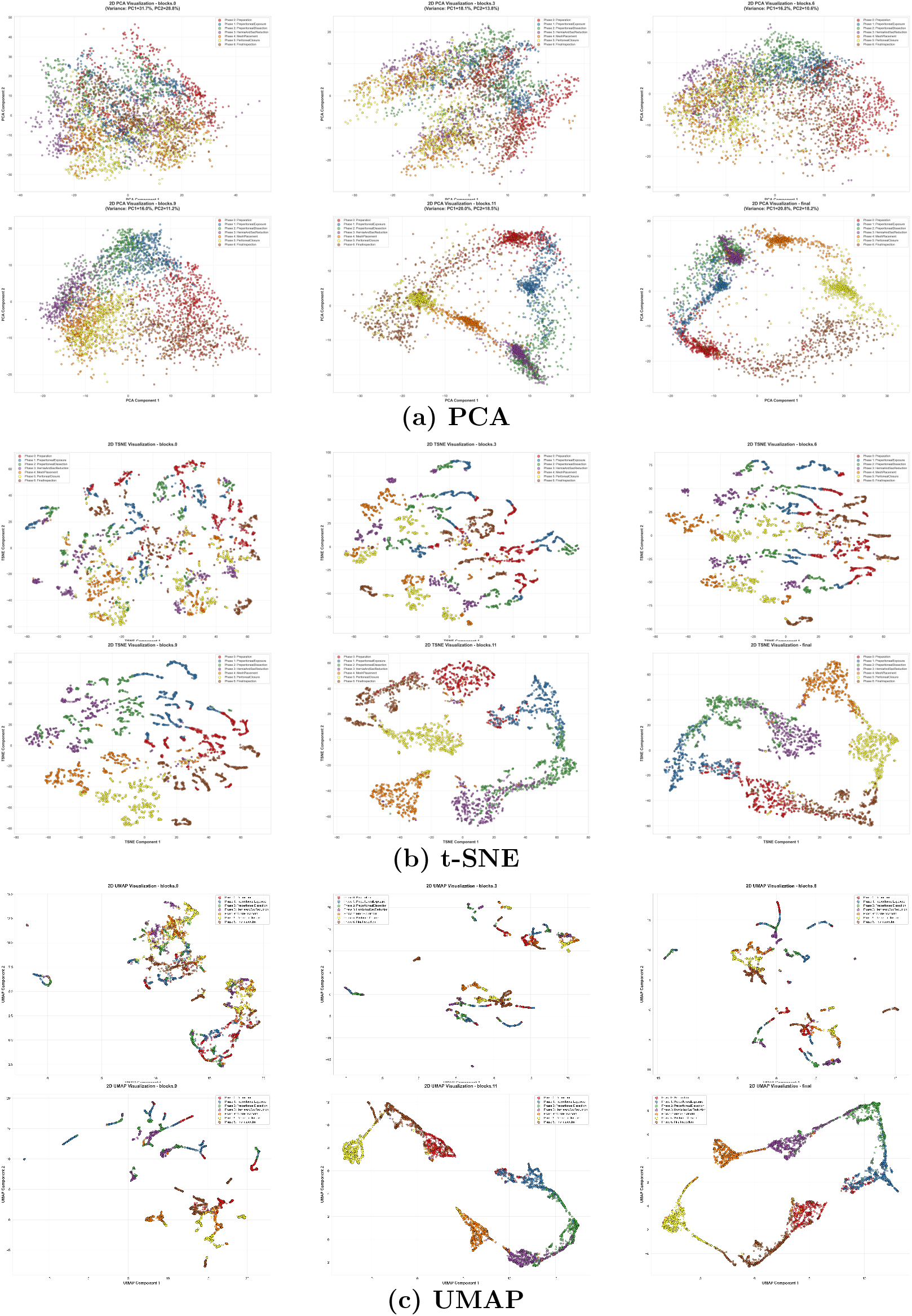
2D Dimensionality Reduction An1a4lysis (PCA, t-SNE, UMAP) of SurgFormer internal feature embeddings across progressive network depths. The visualizations capture the network’s learning dynamics, evolving from highly entangled low-level visual features in Block 0 to distinctly separable, high-level semantic clusters corresponding to the 7 surgical phases in the Final output layer.

**Figure 6:**
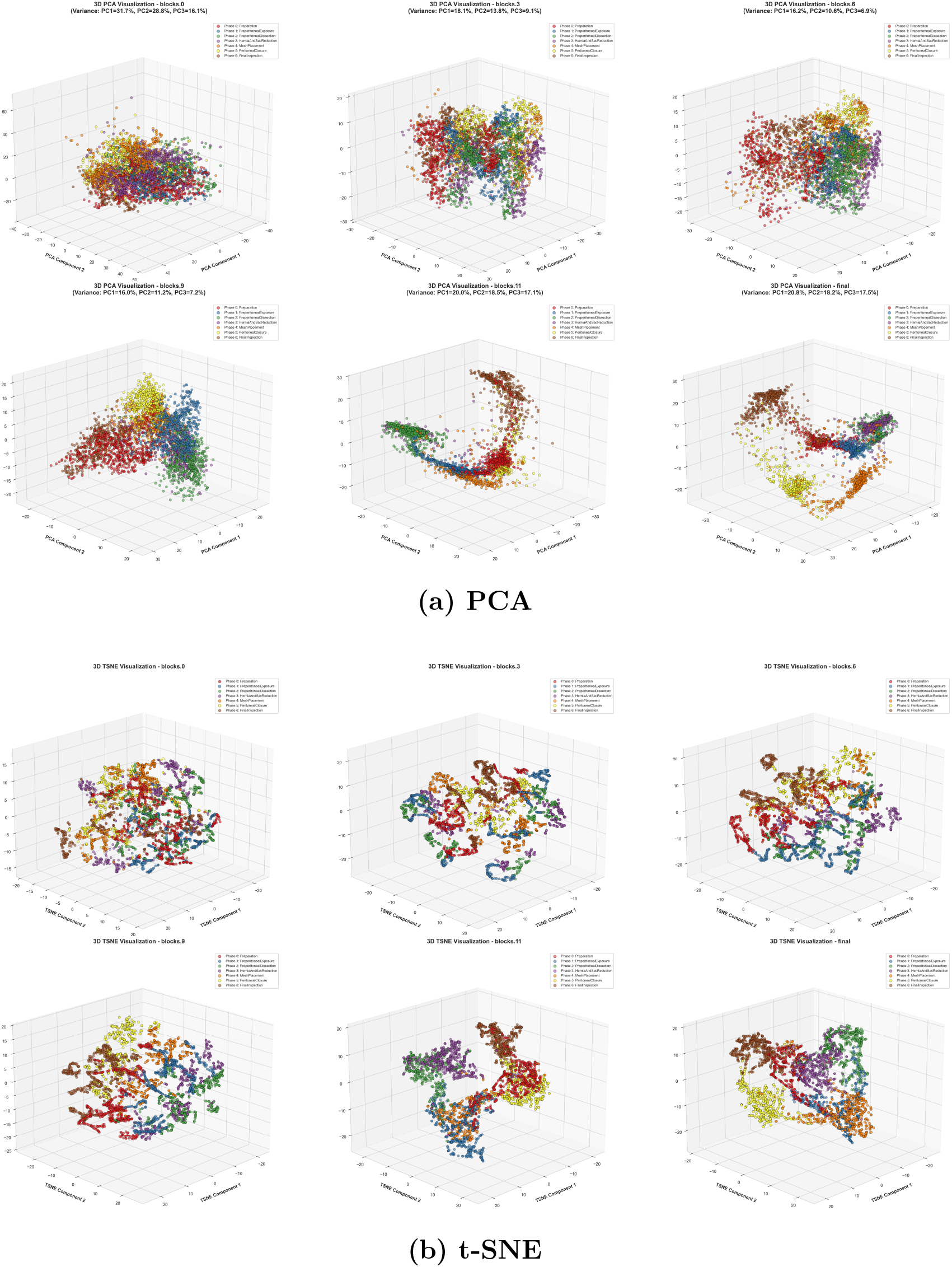
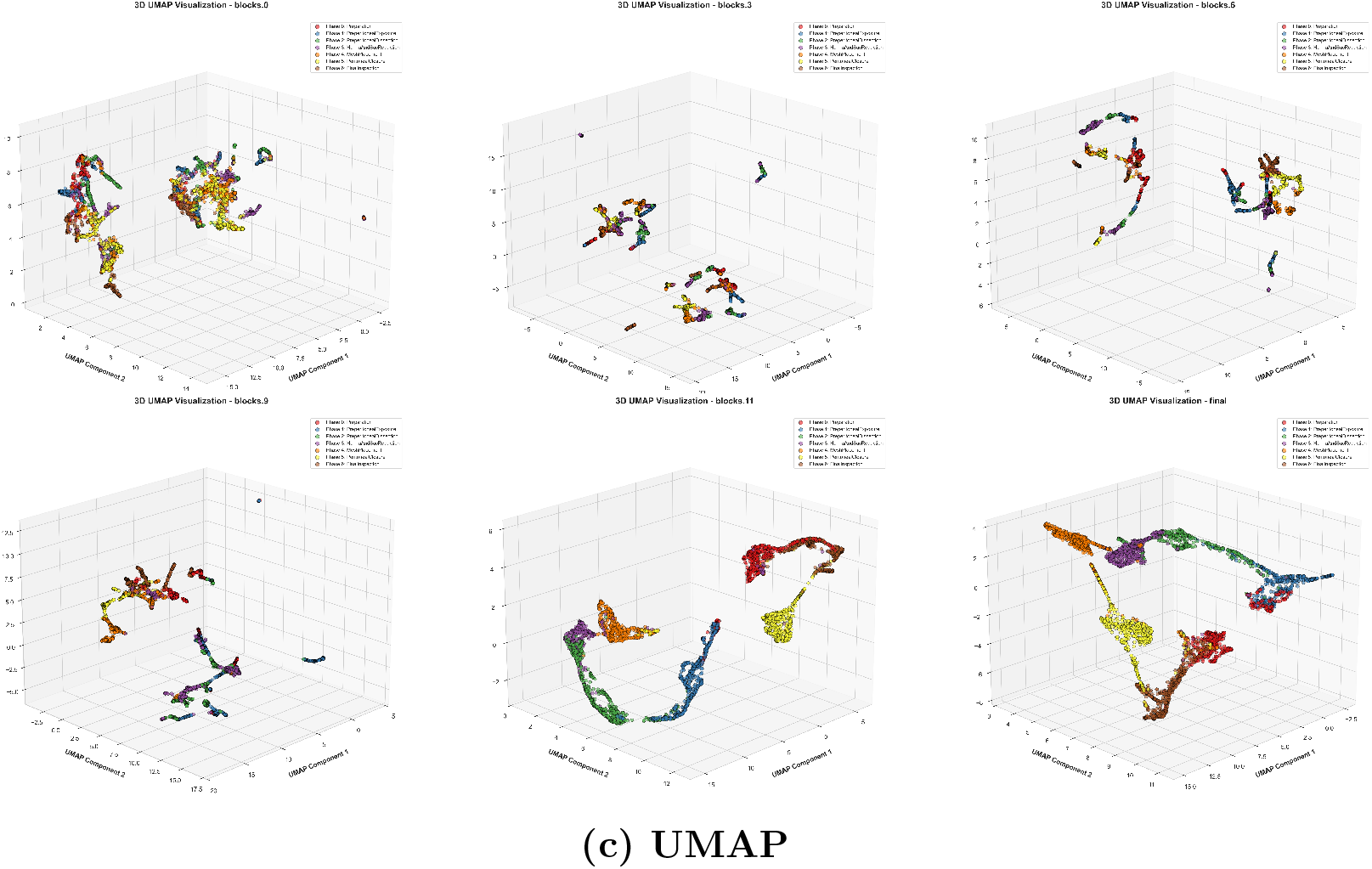
3D Dimensionality Reduction Analysis (PCA, t-SNE, UMAP) of SurgFormer internal feature embeddings across progressive network depths. The visualizations capture the network’s learning dynamics, evolving from highly entangled low-level visual features in Block 0 to distinctly separable, high-level semantic clusters corresponding to the 7 surgical phases in the Final output layer.

## 4. Discussion

The integration of context-aware artificial intelligence into complex procedures like LIHR requires models that are not only highly accurate but also highly interpretable. This study demonstrates that the inherent data scarcity in specialized surgical domains can be successfully overcome through sequential transfer learning, as summarized in Table 2. The necessity of this domain-specific adaptation is heavily underscored by our zero-shot evaluation, where the Cholec80-pretrained model achieved a Top-1 accuracy of only 15.77% on the TAPP dataset. By initializing a vision transformer with weights adapted sequentially from generic human actions (Kinetics-400) to standard laparoscopic scenes (Cholec80), the model ultimately achieved an exceptional 90.64% accuracy on unseen TAPP procedures. The rapid convergence observed at epoch 10 during target task fine-tuning underscores the efficiency of this knowledge transfer, proving that robust spatial-temporal priors drastically reduce the data volume required to map novel, highly complex anatomies.

Detailed class-wise analysis via the normalized confusion matrix (Figure 1) reveals the model’s high reliability in identifying distinct procedural steps, such as *Preperitoneal Exposure* (95.3% accuracy), *Hernia & Sac Reduction* (96.9% accuracy), *Mesh Placement* (92.9%) and *Peritoneal Closure* (95.1%). However, it also highlights specific clinical nuances, notably a drop in accuracy during *Preperitoneal Dissection* (65.3%), where 31.6% of frames were misclassified as *Hernia & Sac Reduction* (Figure 1-e). This quantitative finding aligns seamlessly with the qualitative phase bar analysis (Figure 2), which highlights a known phenomenon in surgical data science: the highest density of predictive errors occurs during phase transitions. Because surgical phases are continuous rather than strictly discrete, the exact boundary between steps (e.g., transitioning from dissection to reduction) is often subjective and visually ambiguous due to shared instruments and similar anatomical views. The real-time video overlays (Figure 3) explicitly visualize these brief, transient deviations, providing a transparent mechanism for surgeons to evaluate the model’s reliability in an intraoperative setting.

The core of our experimental investigation centered on the comparison between direct, cascade, and cumulative training protocols. While direct training achieved a high accuracy of 89.89%, it was consistently outperformed by the three-stage sequential transfer learning utilized in the cumulative protocol. This confirms that pre-training on the Cholec80 benchmark provides essential spatial-temporal priors for laparoscopic scenes that cannot be fully replicated by generic video datasets like Kinetics-400 alone.

The cumulative training results (Table 3) provide a clear map of the model’s data requirements. We observed a significant jump in performance from 81.81% to 87.66% when moving from two to four videos. The peak accuracy of 90.64% at 22 videos suggests a less-is-more phenomenon; after reaching a saturation point of procedural diversity, the addition of further videos may introduce subtle noise that slightly degrades the model’s generalizability.

In contrast, the cascade approach’s lower peak of 87.99% highlights the risks of catastrophic forgetting. When the model is trained on small, sequential chunks without revisiting previous data, it appears to lose specialized features learned early in the sequence. These findings collectively prove that for complex procedures like TAPP, the most effective strategy is a multi-stage sequential transfer learning pipeline combined with parallel exposure to a critical mass of approximately 20-22 procedures.

Finally, the progressive embedding analysis demystifies the black box nature of deep learning in surgery. The sequential PCA, t-SNE, and UMAP visualizations (Figures 5 and 6) explicitly prove that the SurgFormer architecture is genuinely learning to construct high-level, discriminative semantic concepts rather than merely memorizing frame sequences. This level of algorithmic transparency is essential for building clinical trust for eventual deployment in the operating room.

## 5. Conclusion

This study establishes a robust and interpretable framework for automated surgical phase recognition in TAPP laparoscopic inguinal hernia repair by leveraging the SurgFormer vision transformer and an optimized three-stage sequential transfer learning strategy. Our findings demonstrate that a cumulative training approach achieves a peak accuracy of 90.64%, significantly surpassing standard fine-tuning benchmarks. A key contribution of this research is the identification of a “less-is-more” phenomenon in data efficiency, where a curated subset of 22 training videos provided the optimal saturation point for generalizability, while the comparison of training protocols highlighted the necessity of parallel data exposure to mitigate catastrophic forgetting. Furthermore, the integration of real-time visualizations and progressive embedding analyses confirms that the network learns distinct, high-level semantic representations rather than merely memorizing localized visual textures. Future efforts will focus on multi-institutional validation to ensure generalizability across diverse surgical environments and the development of hardware-software interfaces for live intraoperative deployment.

## Data Availability

The institutional dataset contains transabdominal preperitoneal (TAPP) laparoscopic inguinal hernia repair (LIHR) surgical video recordings and is not publicly available due to institutional privacy policies and Research Ethics Board (REB) restrictions at the McGill University Health Centre (MUHC). The Cholec80 benchmark dataset utilized for intermediate domain adaptation is publicly available. All primary performance metrics, statistical analyses, and model interpretability results are fully contained within the manuscript.

https://camma.unistra.fr/datasets/

## Acknowledgments

We acknowledge the Surgical Performance Enhancement and Robotics (SuPER) Centre and the McGill University Health Centre (MUHC) for institutional support. We would also like to thank the Theator platform for providing the cloud-based surgical intelligence and annotation environment utilized in this study. This work was supported in part by the Montreal General Hospital Foundation (MGHF) through the Mimi Dupuis Benjamin Award and the Nesbitt-McMaster Award.

## Author contributions

M.L. conceived the study, developed the methodology and software, conducted experiments and analysis, and drafted the manuscript. L.S.F. and A.H. supervised the work and revised the manuscript.

## Ethics approval and consent to participate

This research study was conducted with the formal approval of the McGill University Health Centre (MUHC) Research Ethics Board (REB), identification number: 2025-10948. Approval was obtained prior to the commencement of data collection for the institutional TAPP dataset. All surgical video recordings utilized in this study were strictly de-identified to protect patient confidentiality and were handled in full compliance with established institutional privacy protocols.

